# Online internal speech decoding from single neurons in a human participant

**DOI:** 10.1101/2022.11.02.22281775

**Authors:** Sarah K. Wandelt, David A. Bjånes, Kelsie Pejsa, Brian Lee, Charles Liu, Richard A. Andersen

## Abstract

Speech brain-machine interfaces (BMI’s) translate brain signals into words or audio outputs, enabling communication for people having lost their speech abilities due to diseases or injury. While important advances in vocalized, attempted, and mimed speech decoding have been achieved, results for internal speech decoding are sparse, and have yet to achieve high functionality. Notably, it is still unclear from which brain areas internal speech can be decoded. In this work, a tetraplegic participant with implanted microelectrode arrays located in the supramarginal gyrus (SMG) and primary somatosensory cortex (S1) performed internal and vocalized speech of six words and two pseudowords. We found robust internal speech decoding from SMG single neuron activity, achieving up to 91% classification accuracy during an online task (chance level 12.5%). Evidence of shared neural representations between internal speech, word reading, and vocalized speech processes were found. SMG represented words in different languages (English/ Spanish) as well as pseudowords, providing evidence for phonetic encoding. Furthermore, our decoder achieved high classification with multiple internal speech strategies (auditory imagination/ visual imagination). Activity in S1 was modulated by vocalized but not internal speech, suggesting no articulator movements of the vocal tract occurred during internal speech production. This works represents the first proof-of-concept for a high-performance internal speech BMI.

## Introduction

Speech is one of the most basic forms of human communication, a natural and intuitive way for humans to express their thoughts and desires. Neurological diseases like amyotrophic lateral sclerosis (ALS) and brain lesions can lead to the loss of this ability. In the most severe cases, patients who experience full body paralysis might be left without any means of communication. Patients with ALS self-report loss of speech as their most serious concern (Hecht et al. 2002). Brain-machine Interfaces (BMIs) are devices offering a promising technological path to bypass neurological impairment by recording neural activity directly from the cortex. BMIs have demonstrated potential to restore independence to tetraplegic participants by reading out movement intentions directly from the brain (Aflalo et al. 2015; Andersen et al. 2014; Andersen, Aflalo, and Kellis 2019; Andersen 2019). Similarly, reading out internal (also reported as inner, imagined, or covert) speech signals could allow the restoration of communication to people who have lost it.

Decoding speech signals directly from the brain presents its own unique challenges. While non-invasive recording methods like functional magnetic imaging (fMRI), electroencephalography (EEG) or magnetoencephalography (MEG) (Dash, Ferrari, and Wang 2020; Dash et al. 2020) are important tools to locate speech and internal speech production, they lack the necessary temporal and spatial resolution, adequate signal-to-noise ratio or portability for building an online speech BMI (Luo, Rabbani, and Crone 2022; Martin et al. 2018; Rabbani, Milsap, and Crone 2019). Intracortical electrophysiological recordings have higher signal-to-noise ratios and excellent temporal resolution (Nicolas-Alonso and Gomez-Gil 2012), and are a more suitable choice for internal speech decoding device.

Invasive speech decoding has predominantly been attempted with electrocorticography (ECoG) (Rabbani, Milsap, and Crone 2019) or stereo-electroencephalographic (sEEG) depth arrays (Herff, Krusienski, and Kubben 2020), as they allow sampling neural activity from different parts of the brain simultaneously. Impressive results in vocalized and attempted speech decoding and reconstruction have been achieved using these techniques (Angrick et al. 2018; Herff et al. 2019; Kellis et al. 2010; Makin, Moses, and Chang 2020; Moses et al. 2021). However, vocalized speech has also been decoded from small-scale microelectrode arrays located in the motor cortex (Stavisky et al. 2019; Wilson et al. 2020) and the supramarginal gyrus (SMG) (Wandelt et al. 2022), demonstrating vocalized speech BMIs can be built using neural signals from localized regions of cortex.

While important advances in vocalized speech (Makin, Moses, and Chang 2020), attempted speech (Moses et al. 2021) and mimed speech (Bocquelet et al. 2016; Anumanchipalli, Chartier, and Chang 2019) decoding have been made, highly accurate internal speech decoding has not been achieved. Lack of behavioral output, lower signal-to-noise ratio, and differences in cortical activation have resulted in much lower classification accuracies of internal speech (Angrick et al. 2018; Martin et al. 2018; Luo, Rabbani, and Crone 2022; Proix et al. 2022). In Pei, Barbour, *et al*., 2011 patients implanted with ECoG grids over frontal, parietal and temporal regions silently read or vocalized written words from a screen. Researchers significantly decoded vowels (37.5%) and consonants (36.3%) from internal speech (chance level 25%). (Ikeda et al. 2014) decoded three internally spoken vowels using ECoG arrays using frequencies in the beta band, with up to 55.6% accuracy from Broca area (chance level 33%). Using the same recording technology, Martin *et al*., 2016 investigated the decoding of six words during internal speech. The authors demonstrated an average pair-wise classification accuracy of 58%, reaching 88% for the highest pair (chance level 50%). These studies were so-called open loop experiments, in which the data was analyzed offline after acquisition. A recent paper demonstrated real-time (closed loop) speech decoding using stereotactic depth electrodes (Angrick et al. 2021). Results were encouraging as internal speech could be detected; however, the reconstructed audio was not discernable and required audible speech to train the decoding model.

While to our knowledge internal speech has not previously been decoded from SMG, evidence for internal speech representation in SMG exists. In a review of 100 fMRI studies, (Cathy J. Price 2010) described SMG activity not only during speech production, but also suggested its involvement in subvocal speech (Langland-Hassan and Vicente 2018; Perrone-Bertolotti et al. 2014). Similarly, an ECoG study identified high frequency SMG modulation during vocalized and internal speech (Pei, Leuthardt, et al. 2011a). Additionally, fMRI studies demonstrated SMG involvement in phonologic processing; for instance, the participant decided if two words rhyme (Oberhuber et al. 2016). Performing such tasks requires the participant to internally “hear” the word, indicating potential internal speech representation (Binder 2017). Furthermore, a study performed in people suffering from aphasia found lesions in SMG and its adjacent white matter affected inner speech rhyming tasks (Geva *et al*., 2011). Recently, (Makin, Moses, and Chang 2020) showed electrode grids over SMG contributed to vocalized speech decoding. Finally, vocalized grasps and color words were decodable from SMG from the same participant involved in this work (Wandelt et al. 2022). These studies provide evidence for the possibility of an internal speech decoder from neural activity in SMG.

The relationship between inner speech and vocalized speech is still debated. The general consensus posits similarities between internal and vocalized speech processes (Pei, Leuthardt, et al. 2011a), but the degree of overlap is not well understood (Cooney, Folli, and Coyle 2022; 2022; Martin et al. 2018; Perrone-Bertolotti et al. 2014; Alderson-Day and Fernyhough 2015). Characterizing similarities between vocalized and internal speech could provide evidence that results found with vocalized speech could translate to internal speech. However, such a relationship may not be guaranteed. For instance, some brain areas involved in vocalized speech might be poor candidates for internal speech decoding.

In this work, a participant with tetraplegia performed internal and vocalized speech of six words and two pseudowords, while neurophysiological responses were captured from two implant sites. We investigated representations of various language processes at the single neuron level using recording microelectrode arrays from the supramarginal gyrus (SMG) located in the posterior parietal cortex (PPC) and the arm region of the primary somatosensory cortex (S1). Words were presented with an auditory or a written cue, and were produced internally as well as orally. We hypothesized SMG and S1 activity would modulate during vocalized speech and SMG activity would modulate during internal speech. Shared representation between internal speech, vocalized speech, auditory comprehension, and word reading processes were investigated.

## Results

### Task design

We characterized neural representations of four different language processes within a population of SMG neurons: auditory comprehension, word reading, internal speech, and vocalized speech production. In this manuscript, internal speech refers to engaging a prompted word internally (“inner monologue”), without correlated motor output, while vocalized speech refers to audibly vocalizing a prompted word.

The task contained six phases: an inter-trial interval (ITI), a cue phase (Cue), a first delay (D1), an internal speech phase (Internal), a second delay (D2), and a vocalized speech (Speech) phase. Words were cued with either an auditory or a written version of the word **(Figure 1A)**. Six of the words were informed by Martin et al., 2016 (Battlefield, Cowboy, Python, Spoon, Swimming, Telephone). Two pseudowords (Nifzig, Bindip) were added to explore phonetic representation in SMG.

**Figure 1.**
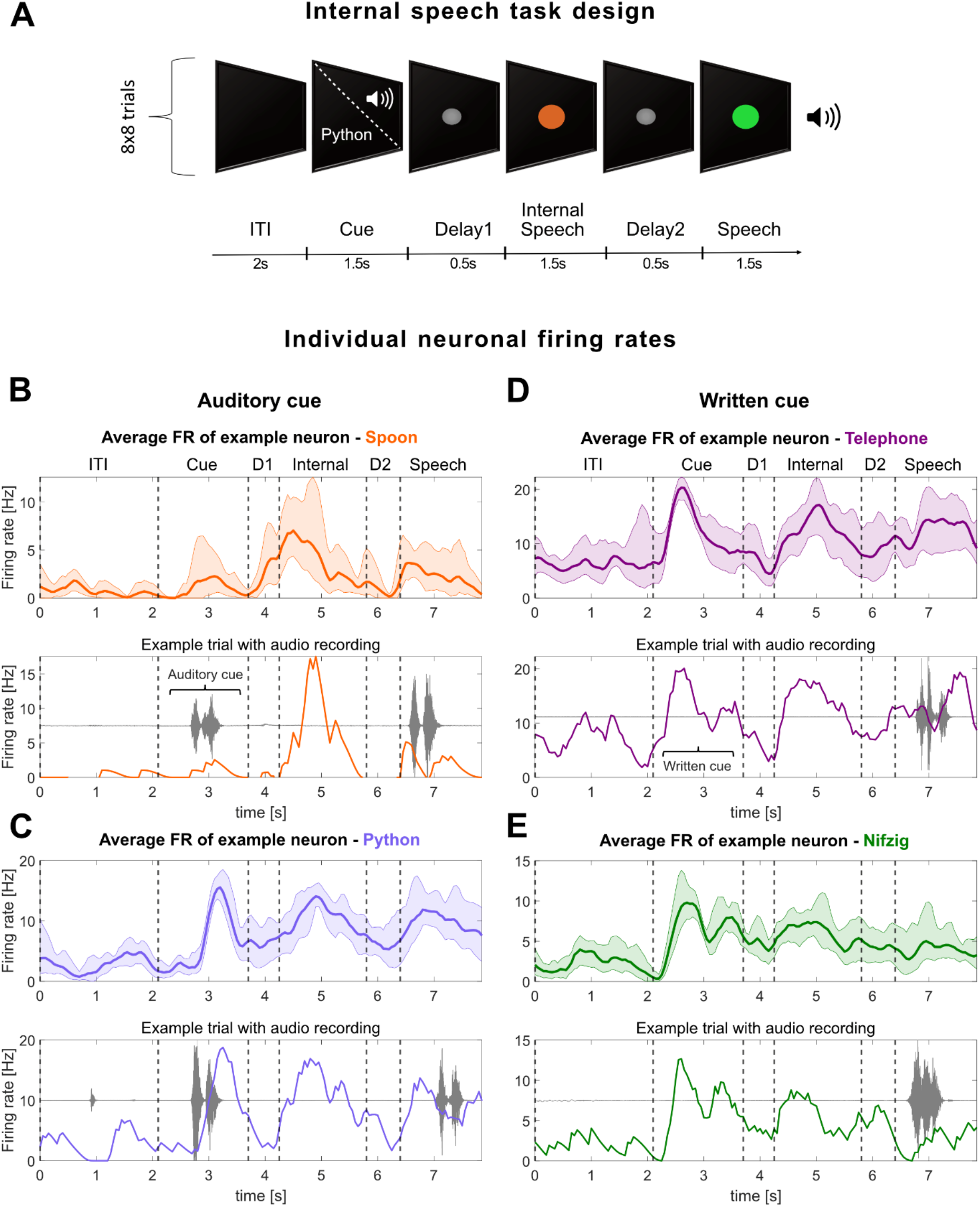
Neurons in the supramarginal gyrus represent language processes. **A)** Written words and sounds were used to cue six words and two pseudowords in a tetraplegic participant. The “Audio cue” task was composed of an inter-trial interval (ITI), a cue phase during which the sound of one of the words was emitted from a speaker [between 842 – 1130ms], a first delay (D1), an internal speech phase, a second delay (D2) and a vocalized speech phase. The “Written cue” task was identical to the “Audio cue” task, apart that written words appeared on the screen for 1.5 seconds. Eight repetitions of eight words were performed per session day and per task. **B,C)** Example smoothed firing rates of neurons tuned to four words in SMG during the “Audio cue” and **D,E)** the “Written cue” task. The top part of each word figure shows the average firing rate over eight trials (solid line: mean, shaded area: 95% bootstrapped confidence interval). The bottom part of each figure shows one out of eight example trials with associated audio amplitude (gray). Vertically dashed lines indicate the beginning of each phase.

### Single neurons modulate firing rate during internal speech in SMG

For each of the four language processes, we observed selective modulation of individual neurons’ firing rates **(Figure 1B,C)**. In general, firing rates of neurons increased during the active phases (Cue, Internal, Speech) and decreased during rest phases (ITI, D1, D2). A variety of activation patterns were present in the neural population. Example neurons were selected to demonstrate increases in firing rates during internal speech **(Figure 1B**, Spoon); neurons were also active during the cue and vocalized speech **(Figure 1B, Figure S1)**. Regardless of the cue modality (auditory in **Figure 1B,C**, written in **Figure 1D,E**), internal speech highly modulated individual neuron firing rates.

These stereotypical activation patterns were evident at the single trial level (**Figure 1A,B,C,D** bottom panel). When the auditory recording was overlaid with firing rates from a single trial, a heterogeneous neural response was observed **(Figure S1A)**, with some SMG neurons preceding or lagging peak auditory levels during vocalized speech. In contrast, neural activity from primary sensory cortex (S1) only modulated during vocalized speech, and produced stereotyped firing patterns regardless of the vocalized word **(Figure S1B)**.

### Population activity represented selective tuning for individual words

Population analysis in SMG mirrored single neuron patterns of activation, showing steep increases in tuning during the active task phases (**Figure 2A**). Tuning of a neuron to a word was determined by fitting a linear regression model to the firing rate in 50 ms time bins (methods). Representation of the auditory cue was lower compared to the written cue **(Figure 2B, Cue)**. However, this difference was not observed for other task phases. The tuned population activity in S1 increased during vocalized speech, but not during the cue and internal speech phases **(Figures S2A)**.

**Figure 2.**
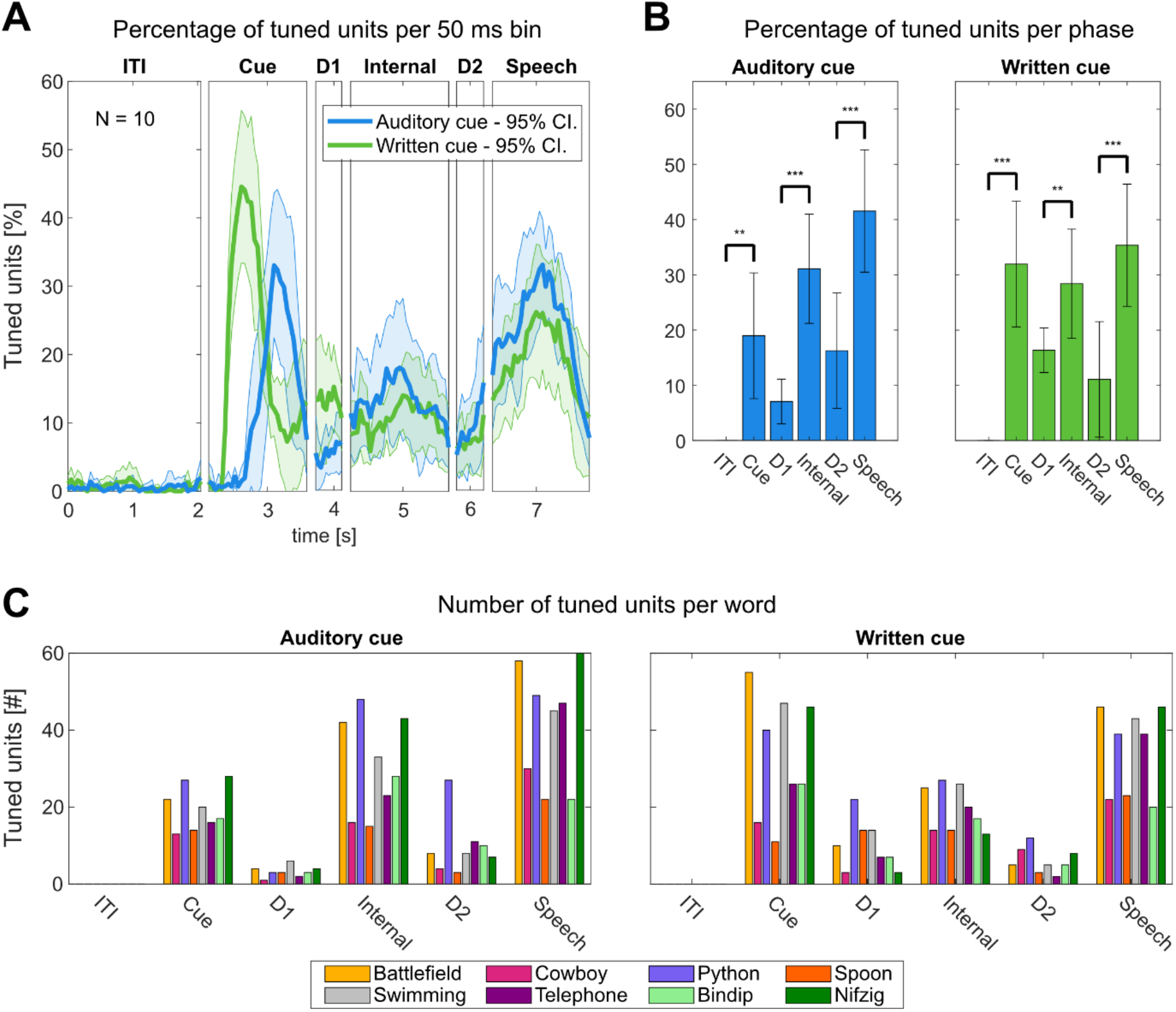
Neuronal population SMG activity represents individual words. **A)** Average percentage of tuned neurons to words in 50ms time bins in SMG over the trial duration for “Auditory cue” (blue) and “Written cue” (green) tasks (solid line: mean over 10 sessions, shaded area: 95% confidence interval). The written cue appeared on average 250ms earlier on the screen than the auditory sound. **B)** Average percentage of tuned neurons computed on firing rates per task phase, with 95% confidence interval over 10 sessions. Tuning during action phase (Cue, Internal, Speech) following rest phases (ITI, D1, D2) was significantly higher (t-test: **p < 0.01, ***p < 0.001). **C)** Number of neurons tuned to each individual word in each phase for the “Auditory cue” and “Written cue” tasks.

To quantitatively compare activity between phases, we computed selectivity for individual words from the average FR in each task phase **(Figure 2B,C)**. Tuning during the cue, internal speech and vocalized speech phases was significantly higher compared to their preceding rest phases ITI, D1, and D2 (t-test, ** = p < 0.01, *** = p < 0.001). Representation for all words was observed in each phase, including pseudowords (Bindip and Nifzig) **(Figure 2C)**.

The neural population in SMG simultaneously represented several distinct aspects of language processing: temporal changes, input modality (auditory, written), and unique words from our vocabulary list **(Figure 3)**. We used demixed Principal Components Analysis (dPCA) to decompose and analyze contributions of each individual component: Timing, Cue Modality, and Word.

**Figure 3.**
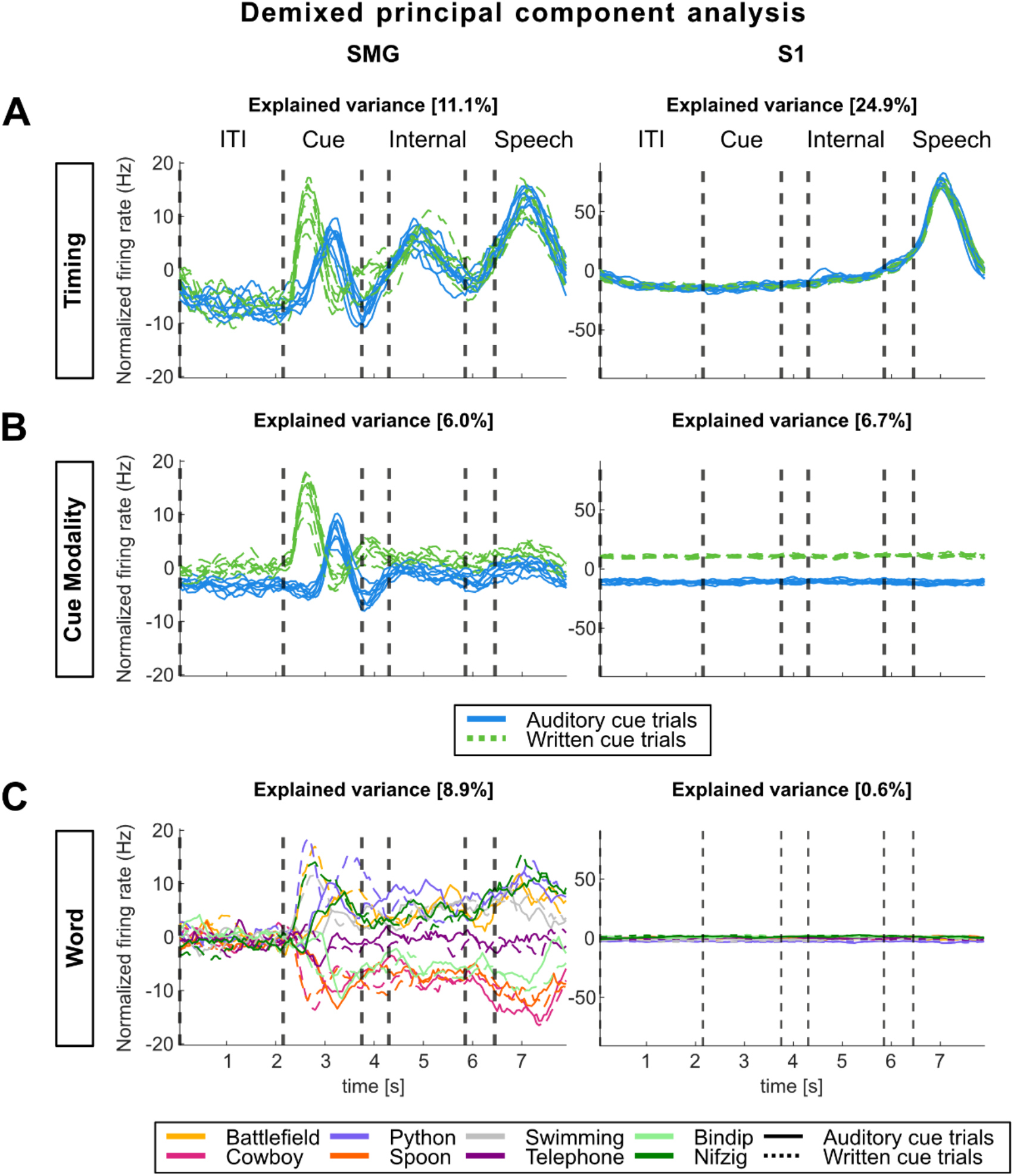
Demixed principal component analysis (dPCA) highlights SMG’s involvement in language processing. dPCA was performed to investigate variance within three marginalizations: “Timing”, “Cue Modality”, and “Word”. Demixed principal components explaining the highest variance within each marginalization were plotted over time. **A)** The “Timing” marginalization demonstrates SMG modulation during cue, internal speech and vocalized speech, while S1 only represents vocalized speech. **B)** The “Cue Modality” marginalization suggests internal and vocalized speech representation in SMG are not affected by the cue modality. **C)** The “Word” marginalization shows high variability for different words in SMG, but near zero for S1.

The “Timing” component revealed temporal dynamics in SMG peaked during all active phases **(Figure 3A)**. In contrast, temporal S1 modulation peaked only during vocalized speech production, indicating a lack of synchronized lip and face movement of the participant during the other task phases **(Figure 3B)**. While “Cue Modality” components were separable during the cue phase, they overlapped during subsequent phases. Thus, internal and vocalized speech representation may not be influenced by the cue modality. Pseudowords had similar separability to lexical words **(Figure 3C)**. The explained variance between words was close to zero in S1.

### Internal speech is highly decodable in SMG

Separable neural representation of both internal and vocalized speech processes implicate SMG as a rich source of neural activity for real-time speech BMI devices. All words in our vocabulary list were highly decodable, averaging 55% offline decoding and 84% online decoding from neurons during internal speech (Figure 4AB). Words spoken during the vocalized phase were also highly discriminable, averaging 74% offline (Figure 4A).

**Figure 4.**
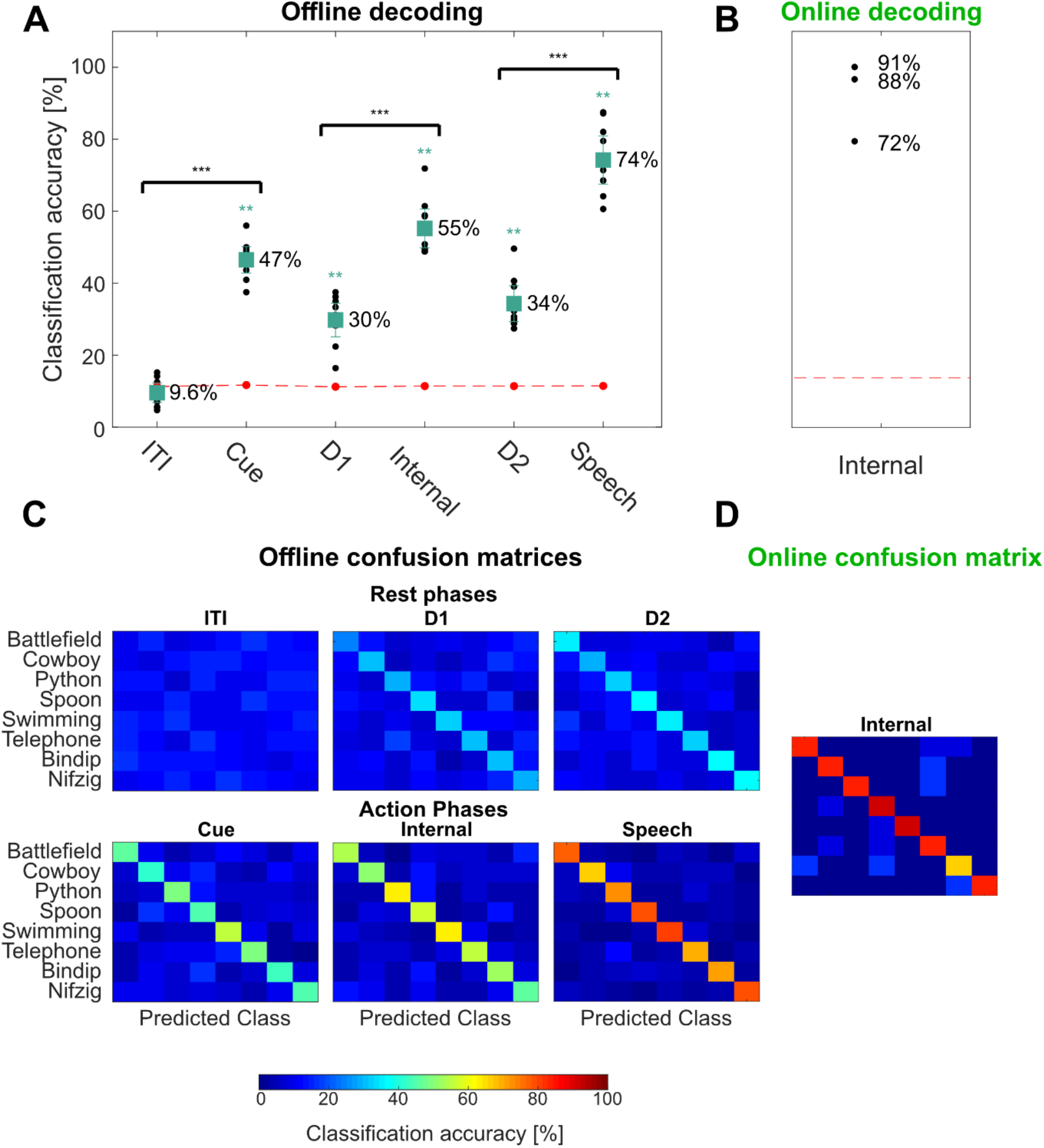
Words can significantly be decoded during internal speech in SMG. **A)** Offline decoding accuracies: “Audio cue” and “Written cue” tasks data were combined for each individual session day, and leave one out cross-validation was performed (black dots). PCA was performed on the training data, a LDA model was constructed, and classification accuracies were plotted with 95% c.i, over the session means. Significance of classification accuracies was evaluated by comparing results to a shuffled distribution (averaged shuffle results = red dots, * = p < 0.05, ** = p < 0.01). Classification accuracies during action phases (Cue, Internal, Speech) following rest phases (ITI, D1, D2) were significantly higher (t-test: ***p < 0.001). **B)** Online decoding accuracies: Classification accuracies for internal speech were evaluated in a closed-loop internal speech BMI application. 72%, 88% and 91% classification accuracies were achieved using respectively 8, 12 and 16 trials per word to train the classification model. **C)** Offline confusion matrix: Confusion matrices for each of the different task phases were computed on the tested data, and averaged over all session days. **D)** Online confusion matrix: A confusion matrix was computed combing the three online runs.

For offline analysis, trial data from both types of cues (auditory and written) were concatenated, since SMG activity was only differentiable between the type of cue during the cue phase (**Figure 2A, Figure 3B)**. This resulted in 16 trials per condition. Features were selected via principal component analysis (PCA) on the training dataset, and principal components (PCs) which explained 95% of the variance were kept. A linear discriminant analysis (LDA) model was evaluated with leave-one-out cross-validation (CV). Significance was computed by comparing results to a null distribution (methods).

Significant word decoding was observed during all phases, except during the ITI (**Figure 4A**, null distribution, turquoise - p < 0.001 = ***). Decoding accuracies were significantly higher in the cue, internal speech, and speech condition, compared to rest phases ITI, D1 and D2 (**Figure 4A**, t-test, black – p < 0.001 = ***). Significant cue phase decoding suggested that modality-independent linguistic representations were present early within the task (Leuthardt et al. 2012). Internal speech decoding averaged 55% offline, with the highest session at 72% and a chance level of ∼12.5% (**Figure 4A**, red line). Vocalized speech averaged even higher, at 74%. All words were highly decodable (**Figure 4C**). As suggested from our dPCA results, individual words were not significantly decodable from neural activity in S1 (**Figure S2B**), indicating generalized activity for vocalized speech in the S1 arm region **(Figure 3C)**.

### High accuracy online speech decoder

We developed an online, closed-loop internal speech BMI with the highest achievable accuracy of 91% in an eight-word vocabulary **(Figure 4B)**. A training dataset was generated using the written cue task, with 8 repetitions of each word. An LDA model was trained on the internal speech data of the training set, corresponding to only 96 seconds of neural data. The trained decoder predicted internal speech during the online task. During the online task, the vocalized speech phase was replaced with a feedback phase. The decoded word was shown in green if correctly decoded, and in red if wrongly decoded **(Video S1)**.

The classifier was retrained after each run of the online task, adding the newly recorded data. The first online run was trained on 8 repetitions per word and achieved 72% internal speech decoding. The second was trained on 12 trial repetitions and achieved 88% classification accuracy, while the third run was trained on 16 trial repetitions and achieved 91% classification accuracy. All words were well represented, illustrated by a confusion matrix (Figure 4D).

### Shared representations between internal speech, written words, and vocalized speech

Several different language processes are engaged during the task; auditory comprehension or visual word recognition during the cue phase, and internal speech and vocalized speech production during the speech phases. It has been widely assumed each of these processes are part of a highly distributed network, involving multiple cortical areas (Indefrey and Levelt 2004). In this work, we observed significant representation of each of these processes in a common cortical region, SMG. To explore the relationships between each of these processes, we used cross-phase classification to identify the distinct and common neural codes. By training our classifier on the representation found in one phase (e.g. the cue phase) and testing the classifier on another phase (e.g. internal speech), we quantified generalizability of our models across neural activity of different language processes **(Figure 5)**. The generalizability of a model to different task phases was evaluated through a t-test. No significant difference between classification accuracies indicated good generalization of the model, while significantly lower classification accuracies suggested poor generalization of the model.

**Figure 5.**
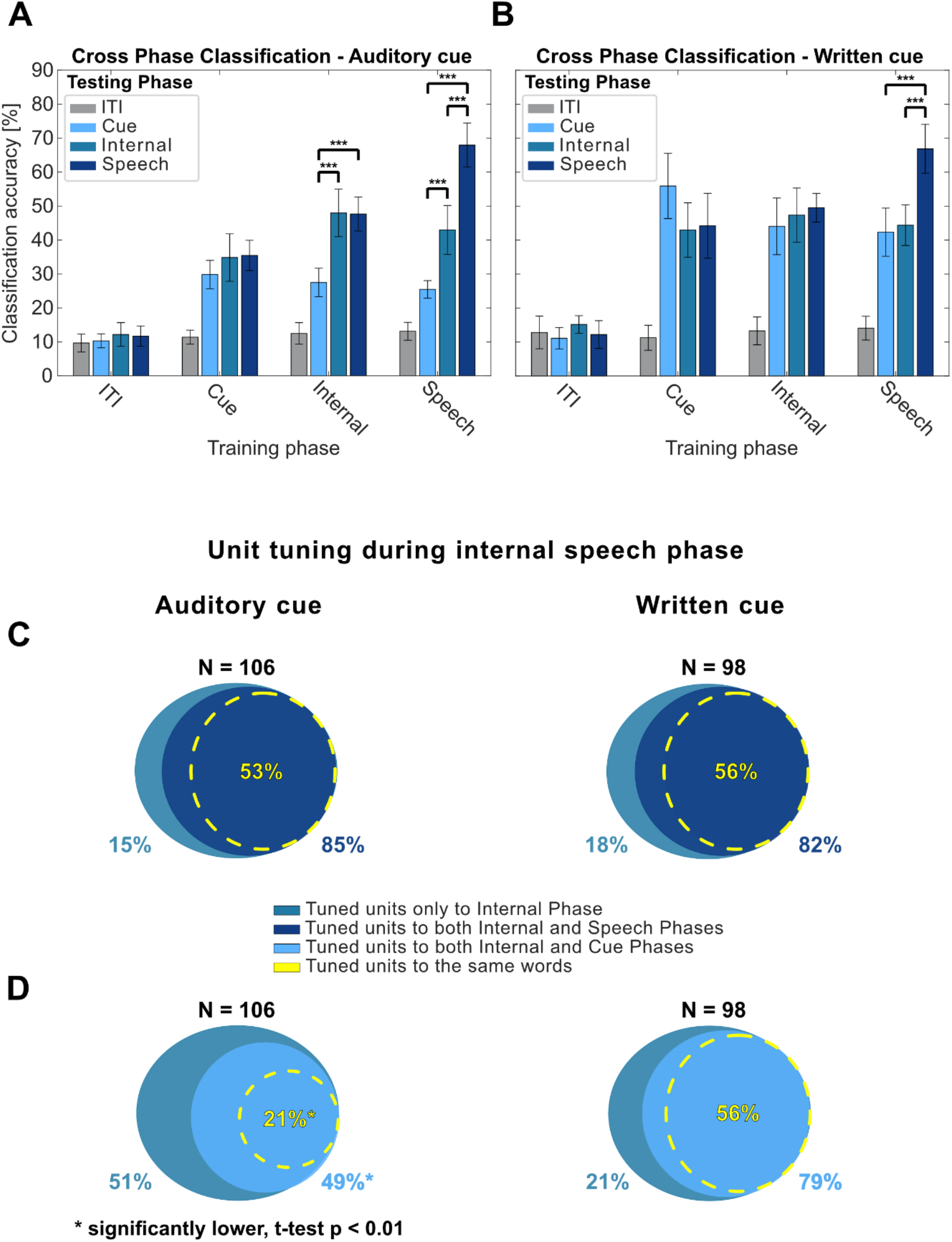
Shared representations between internal speech, vocalized speech, and written word processing. **A)** Evaluating the overlap of shared information between different task phases in the “Auditory cue” task. For each of the 10 session days, cross-phase classification was performed. It consisted in training a model on a subset of data from one phase (e.g. Cue) and applying it on a subset of data from ITI, cue, internal and speech phases. This analysis was performed separately for each task phase. PCA was performed on the training data, a LDA model was constructed, and classification accuracies were plotted with a 95% c.i over session means. Significant differences in performance between phases were evaluated between the 10 sessions (t-test, p < 0.001). For easier visibility, significant differences between ITI and other phases were not plotted. **B)** Same as A) for “Written cue” task. **C)** Percentage of neurons tuned during the internal speech phase which are also tuned during the vocalized speech phase. Neurons tuned during the internal speech phase were computed as in Figure 2B separately for each session day. From these, the percentage of neurons that was also tuned during vocalized speech was calculated. More than 80% of neurons during internal speech were also tuned during vocalized speech (85% in the “Auditory cue” task, 82% in the “Written cue” task). 53% of “Auditory cue” and 56% “Written cue” neurons also showed tuning to the same words during internal speech and vocalized speech phases. **D)** Percentage of neurons tuned during the internal speech phase which are also tuned during the cue phase. 79% of neurons tuned during internal speech were also tuned during the written cue phase (right side). A smaller 49% of neurons tuned during the internal speech phase were also tuned during the auditory cue phase. Percentages between Internal-Speech (85%,82%) and Internal-Written cue (79%) were not significantly different. However, the overlapping Internal-Auditory cue percentage (49%) was significantly lower compared to the other overlaps (pairwise t-test, p < 0.01 for each comparison). 56% of neurons were tuned to the same words during the written cue phase and the internal speech phase. A significantly lower 21% of neurons were tuned to the same words during the auditory cue and the internal speech phase (pairwise t-test, p < 0.01 for each comparison).

The strongest shared neural representations were found between visual word recognition, internal speech, and vocalized speech **(Figure 5B)**. A model trained on internal speech was highly generalizable to both vocalized speech and written cued words, evidence for a possible shared neural code (**Figure 5B**, Internal). In contrast, the model’s performance was significantly lower when tested on data recorded in the auditory cue phase (**Figure 5A**, Internal, t-test, p < 0.001).

Additionally, a model trained on vocalized speech generalized equally well to internal speech and written words (**Figure 5B**, Speech). However, the model generalized significant better to internal speech than the representation found during the auditory cue (**Figure 5A**, Speech, Internal vs Auditory Cue).

Neuronal representation of words at the single neuron level was highly consistent between internal speech, vocalized speech, and written cue phases. A high percentage of neurons were not only active during the same task phases, but also were tuned to the same words (**Figure 5C,D**). 82-85% of neurons active during internal speech were also active during vocalized speech. In 53-56% of neurons, tuning was preserved between the internal speech and vocalized speech phases **(Figure 5C)**. During the cue phase, 79% of neurons active during internal speech were also active during the written cue (**Figure 5D**, right). However, a significantly lower percentage of neurons (49%, t-test, p < 0.01 for all three other comparisons) were active during the auditory cue phase (**Figure 5D**, left). Similarly, 56% of neurons were tuned to the same words during the written cue phase as during the internal speech phase, while a significantly lower 21% of neurons were tuned to the same words during the auditory cue phase as during the internal speech phase.

Together with the cross-phase analysis, these results suggest strong shared neural representations between internal speech, vocalized speech, and written cue phase, both at the single neuron and population level.

### Robust decoding of multiple internal speech strategies within SMG

This shared neural representation between written, inner, and vocalized speech suggests that all three partly represent the same cognitive process or all cognitive processes share common neural features. While internal and vocalized speech have been shown to share common neural features (Pei, Leuthardt, et al. 2011a), similarities between internal speech and the written cue could have occurred through several different cognitive processes. For instance, the participant’s observation of the written cue could have activated silent reading. This process has been self-reported as activating internal speech, which can involve “hearing” a voice, thus having an auditory component (Alderson-Day and Fernyhough 2015; Alderson-Day, Bernini, and Fernyhough 2017). However, the participant could also have mentally pictured an image of the written word while performing internal speech, involving visual imagination in addition to language processes. Both hypotheses could explain the high amount of shared neural representation between the written cue and the internal speech phases **(Figure 5B)**.

We therefore compared two possible internal sensory strategies: a “sound imagination” strategy in which the participant imagined hearing the word, and a “visual imagination” strategy in which the participant visualized the picture of the word **(Figure S3A)**. Both strategies were cued by each modality tested previously (auditory and written words).

Both employed strategies highly represented the four-word dataset (**Figure S3B**, highest 94%, chance level: 25%). Furthermore, the participant described the “sound imagination” strategy as being easier and more similar to the internal speech condition of the first experiment. The participant’s self-reported strategy suggests no visual imagination was performed during internal speech. Correspondingly, similarities between written cue and internal speech phases may stem from internal speech activation during the silent reading of the cue. These results suggest our speech BMI decoder is robust to multiple types of internal speech strategies.

### Evidence of phonetic representation in SMG

Shared neural representations of the observed activation patterns could also have occurred if SMG encoded the semantic content of words. The native bilingual participant translated a four-word English vocabulary into Spanish, ensuring the semantic meaning of each word remained the same, while changing the phonetic content **(Figure S4A)**. Then, an English version of the task and a Spanish version of the task were run on three separate session days. Data were aggregated over languages and days, a support vector machine classifier model was used to evaluate the testing set with 8-fold cross-validation, and a confusion matrix was calculated. Both English and Spanish words were highly represented and separable. The offline model rarely confused words with the same semantic meaning, suggesting strong neural representation of phonetic content in SMG.

## Discussion

In this work, we demonstrated a robust decoder for internal and vocalized speech, capturing single neuron activity from the supramarginal gyrus. A chronically implanted, speech-abled participant with tetraplegia was able to use an online, closed-loop internal speech BMI to achieve up to 91% classification accuracy with an eight-word vocabulary. Furthermore, high decoding required only 24 seconds of training data per word. Firing rates recorded from S1 showed generalized activation only during vocalized speech activity, but individual words were not classifiable. In SMG, shared neural representations between internal speech, the written cue, and vocalized speech suggest the occurrence of common processes. Robust control could be achieved using multiple internal speech strategies and for both English and Spanish vocabularies. Representation of pseudowords provided evidence for phonetic word encoding in SMG.

### Single neurons in the supramarginal gyrus encode internal speech

We demonstrated internal speech decoding of six different words and two pseudowords in SMG. Single neurons increased their firing rates during internal speech **(Figure 1, S1)**, which was also reflected at the population level **(Figure 2A,B)**. Each word was well represented in the neuronal population **(Figure 2C, 3C**). Classification accuracy and tuning during the internal speech phase were significantly higher than during the previous delay phase **(Figure 2B, Figure 4A)**. This evidence suggests we did not simply decode sustained activity from the cue phase, but activity generated by the participant performing internal speech. We obtained up to 72% offline classification accuracy of internal speech, and up to 91% during closed-loop online experiments **(Figure 4A)**. These findings provide strong evidence for internal speech processing at the single neuron level in SMG.

### Neurons in primary somatosensory cortex are modulated by vocalized but not internal speech

Neural activity recorded from S1 served as a control for synchronized face and lip movements during internal speech. While vocalized speech robustly activated sensory neurons, no increase of baseline activity was observed during the internal speech phase or the auditory and written cue phases **(Figure 3, S1)**. These results underline no synchronized movement inflated our decoding accuracy of internal speech (**Figure S2**).

A previous imaging study achieved significant decoding of several different internal speech sentences performed by patients with mild ALS (Dash et al. 2020). Together with our findings, these results suggest a BMI speech decoder which does not rely on any movement may translate to communication opportunities for patients suffering from ALS and locked-in syndrome.

### Different face activities are observable but not decodable in arm area of S1

The topographic representation of body parts in S1 has recently been found to be less rigid than previously thought. Generalized finger representation was found in a presumably S1 arm region of interest (ROI) (Rosenthal et al. 2022). Furthermore, a fMRI paper found observable face and lip activity in S1 leg and hand ROIs. However, differentiation between two lip actions was restricted to the face ROI (Muret et al. 2022). Correspondingly, we observed generalized face and lip activity in an predominantly S1 arm region (see Armenta Salas *et al*., 2018 for implant location) during vocalized speech **(Figure 3A, Figure S1, Figure S2A)**. Recorded neural activity contained similar representations for different spoke words **(Figure 3C)** and was not significantly decodable **(Figure S2B)**.

### Shared neural representations between internal and vocalized speech

The extent to which internal and vocalized speech generalize is still debated (Cooney, Folli, and Coyle 2018; Perrone-Bertolotti et al. 2014; Alderson-Day and Fernyhough 2015), and depends on the investigated brain area (Pei, Leuthardt, et al. 2011b; Soroush et al. 2022). In this work, we found on average stronger representation for vocalized (74%) than internal speech (**Figure 4A**, 55%). Additionally, cross-phase decoding of vocalized speech from models trained on data during internal speech resulted in comparable classification accuracies to those of internal speech (**Figure 5A,B**, Internal). However, in certain contexts we observed identical or better classification accuracy during internal speech than vocalized speech (**Figure 4A**, Online decoding). Better decoding of individual internal words than vocalized words was also observed during the phonetic task (**Figure S4**, Cowboy, Python). Most neurons tuned during internal speech were also tuned to the same words during vocalized speech (53-56%, **Figure 5C**). However, some neurons were only tuned during internal speech, or to different words. These observations also applied to firing rates of individual neurons. Here, we observed neurons that had higher peak rates during the internal speech phase than the vocalized speech phase (**Figure 1**: Spoon, Nifzig; **Figure S1:** Swimming, Cowboy). Together, these results suggest SMG does not present a strict nested hierarchy of internal and imagined speech, where all channels involved during imagined speech are a subset of channels involved during vocalized speech (Soroush et al. 2022).

Similar observations were made when comparing internal speech processes to visual word processes. 79% of neurons were active both in the internal speech phase and the written cue phase, and 56% preserved the same tuning (**Figure 5D**, Written Cue). Additionally, high cross-decoding between both phases was observed **(Figure 5B)**.

### Shared representation between speech and written cue presentation

Observation of a written cue may engage a variety of cognitive processes, such as visual feature recognition, semantic understanding and/or related language processes, many of which modulate similar cortical regions as speech (Leuthardt et al. 2012). Studies have found silent reading can evoke internal speech; it can be modulated by presumed author’s speaking speed, voice familiarity or regional accents (Perrone-Bertolotti et al. 2014; Alderson-Day and Fernyhough 2015; Alderson-Day, Bernini, and Fernyhough 2017; Alexander and Nygaard 2008; Filik and Barber 2011; Lœvenbruck et al. 2005). During silent reading of a cued sentence with a neutral vs increased prosody (madeleine brought me vs. MADELEINE brought me), one study in particular found increased left SMG activation correlated with the intensity of the produced inner speech (Lœvenbruck et al. 2005).

Our data demonstrated high cross-phase decoding accuracies between both written cue and speech phases **(Figure 5B)**. Due to substantial shared neural representation, we hypothesize the participant’s silent reading during the presentation of the written cue may have engaged internal speech processes. However, this same shared representation could have occurred if visual processes were activated in the internal speech phase. For instance, the participant could have performed mental visualization of the written word instead of generating an internal monologue, as the subjective perception of internal speech may vary between individuals (Alderson-Day and Fernyhough 2015; Cooney, Folli, and Coyle 2022)

### Investigating internal speech strategies

In a separate experiment, the participant was prompted to execute different mental strategies during the internal speech phase, consisting of “sound imagination” or “visual word imagination” **(Figure S3A)**. We found robust decoding during the internal strategy phase, regardless which mental strategy was performed **(Figure S3B)**. The participant reported the sound strategy was easier to execute than the visual strategy. Furthermore, the participant reported the sound strategy was more similar to the internal speech strategy employed in prior experiments. This self-report suggests the patient did not perform visual imagination during the internal speech task. Therefore, shared neural representation between internal and written word phases during the internal speech task may stem from silent reading of the written cue. Since multiple internal mental strategies are decodable from SMG, future patients could have flexibility with their preferred strategy. For instance, people with a strong visual imagination may prefer performing visual word imagination.

### Phonetic processing in SMG

Since substantial shared neuronal representation was observed across the different task phases, semantic information in SMG might be responsible. Our native bilingual participant performed the same internal speech experiment using a four-word English and four-word Spanish vocabulary of words with the same meaning. If semantics were encoded, we hypothesized a classification model trained on English and Spanish words would confuse words of the same meaning. Instead, we observed stronger representation of phonetic information, including robust representation of pseudowords with no semantic content **(Figure 2C, Figure 3C, Figure S4)**. Our findings in SMG agree with previous literature reports of stronger SMG representation for phonetic rather than semantic decisions on words (Oberhuber et al. 2016; C. J. Price et al. 1997; Seghier et al. 2004; Sliwinska et al. 2012).

### Audio contamination in decoding result

Prior studies examining neural representation of attempted or vocalized speech must potentially mitigate acoustic contamination of electrophysiological brain signals during speech production (Roussel et al. 2020). During internal speech production, no detectable audio was captured by the audio equipment or noticed by the researchers in the room. In the rare cases the participant spoke during internal speech (3 trials), the trial was removed. Furthermore, if audio had contaminated the neural data during the auditory cue or vocalized speech, we would have likely observed significant decoding in all channels. However, no significant classification was detected in S1 channels. We therefore conclude acoustic contamination did not artificially inflate observed classification accuracies during vocalized speech in SMG.

### Impact on BMI applications

In this work, an online internal speech BMI achieved high-performance from single neuron activity in SMG. The online decoders were trained on as little as eight repetitions of 1.5 seconds per word, demonstrating meaningful classification accuracies can be obtained with merely a few minutes’ worth of training data per day. This proof-of-concept suggests SMG may be able to represent a much larger internal vocabulary. By building models on internal speech directly, our results may translate to people who cannot vocalize speech or are completely locked in.

To summarize, we demonstrate SMG as a promising candidate to build an internal brain-machine speech device. Different internal speech strategies were decodable from SMG, allowing patients to use the methods and languages with which they are most comfortable. We found evidence for phonetic representation during internal and vocalized speech. Adding to previous findings indicating grasp decoding in SMG (Wandelt et al. 2022), we pose SMG as a multipurpose brain-machine interface area.

## Data Availability

All data produced in the present study are available upon reasonable request to the authors

## Acknowledgements

We wish to thank L. Bashford, and I. Rosenthal for helpful discussions and data collection. We wish to thank our study participant FG for his dedication to the study which made this work possible. This research was supported by the NIH National Institute of Neurological Disorders and Stroke Grant U01: U01NS098975 and U01: U01NS123127 (S.K.W, D.B., K.P., C.L. and R.A.A.), and by the T&C Chen Brain-machine Interface center (S.K.W., D.B., R.A.A.).

## Author contributions

S.K.W., D.B. and R.A.A. designed the study. S.K.W. developed the experimental tasks and analyzed the results. S.K.W., D.B. and R.A.A. interpreted the results and wrote the paper. K.P. coordinated regulatory requirements of clinical trials. C.L. and B.L. performed the surgery to implant the recording arrays.

## Methods

### Experimental model and subject details

A tetraplegic participant was recruited for an IRB- and FDA-approved clinical trial of a brain-machine interface and he gave informed consent to participate. The participant suffered a spinal cord injury at cervical level C5 two years prior to participating in the study.

### Method details

#### Implants

The targeted areas for implant were the supramarginal gyrus (SMG), and primary somatosensory cortex (S1) and the left ventral premotor cortex (PMv). In this study, SMG and S1 data was considered. For description of localization fMRI tasks and implant locations see (Armenta Salas et al. 2018). In November 2016, the participant underwent surgery to implant one 96-channel multi-electrode array (Neuroport Array, Blackrock Microsystems, Salt Lake City, UT) in SMG and PMv each, and two 7 × 7 sputtered iridium oxide film - tipped microelectrode arrays with 48 channels each in S1. Data were collected between July 2021 and August 2022.

#### Data collection

Recording began two weeks after surgery and continued one to three times per week. Data for this work were collected between 2021 and 2022. Broadband electrical activity was recorded from the NeuroPort arrays using Neural Signal Processors (Blackrock Microsystems, Salt Lake City, UT). Analog signals were amplified, bandpass filtered (0.3-7500 Hz), and digitized at 30,000 samples/sec. To identify putative action potentials, these broadband data were bandpass filtered (250-5000 Hz), and thresholded at -4.5 the estimated root-mean-square voltage of the noise. For some of the analyses, waveforms captured at these threshold crossings were then spike sorted by manually assigning each observation to a putative single neuron, for others, multiunit activity was considered. On average, 33 sorted SMG units (between 22 – 56) and 83 sorted S1 units (between 59 – 96) were included in the analysis. Auditory data was recorded at 30k Hz simultaneously to the neural data. Background noise was reduced post-recording by using the noise reduction function of the program “Audible”.

#### Experimental Tasks

We implemented different tasks to study language processes in the SMG. The tasks cued six words informed by Martin et al., 2016 (Spoon, Python, Battlefield, Cowboy, Swimming, Telephone) as well as two pseudowords (Bindip, Nifzig). The participant was situated 1m in front of a LED screen (1190 mm screen diagonal), where the task was visualized. The task was implemented using the Psychophysics Toolbox (Brainard, 1997; Pelli, 1997; Kleiner et al, 2007) extension for MATLAB.

### Auditory cue task

Each trial consisted of six phases, referred to in this paper as ITI, Cue, D1 (delay 1), Internal, D2 (delay 2), and Speech. The trial began with a brief inter-trial interval (2 sec), followed by a 1.5s long cue phase. During the cue phase, a speaker emitted the sound of one of the eights words (e.g. Python). Word duration varied between 842 and 1130 ms. Then, after a delay period (gray circle on screen; 0.5s), the participant was instructed to internally say the cued word (orange circle on screen; 1.5 sec). After a second delay (gray circle on screen; 0.5s), the participant vocalized the word (green circle on screen, 1.5 seconds).

### Written cue task

The task was identical to the Auditory cue task, except words were cued in writing instead of audio. The written word appeared on the screen for 1.5 seconds during the cue phase. The auditory cue was played ∼ 250ms later than the written cue appeared on the screen, due to software constraints.

The auditory cue task and written cue task were recorded on 10 different session days.

### Control experiments

Two experiments were run to investigate internal strategies and phonetic processing.

### Internal strategy task

The task was designed to vary the internal strategy employed by the participant during the internal speech phase. Two internal strategies were tested: a sound imagination and a visual imagination. For the “sound imagination” strategy, the participant was instructed to imagine what the sound of the word sounded like. For the “visual imagination” strategy, the participant was instructed to perform mental visualization of the written word. We also tested if the cue modality (auditory or written) influenced the internal strategy. A subset of four words were used for this experiment. This led to four different variations of the task:

| Task variation | Cue modality | Internal strategy |
| --- | --- | --- |
| Auditory - Sound | Auditory | Sound imagination |
| Written – Visual | Written | Visual imagination |
| Auditory – Visual | Auditory | Visual imagination |
| Written – Sound | Written | Sound imagination |

### English/Spanish task

The task was designed to understand if SMG stronger encodes semantic or phonetic information. For this task, we made use of the participant’s bilingual abilities and asked him to translate a subset of the words into their Spanish counterparts. This ensured the semantic meaning of words remained the same, while varying the phonetic content. Those words were used to design the Spanish version of the task. On each session day, an English and a Spanish version of the task was run.

The internal strategy task was run on one session day, and the semantic – phonetic task was run on three session days.

### Online task

The “Written cue task” was turned into a closed-loop experiment. To obtain training data for the online task, a written cue task was run. Then, a classification model was trained only on the internal speech data of the task (see classification subsection). The closed-loop task was nearly identical to the “Written cue task” but replaced the vocalized speech phase by a feedback phase. Feedback was provided by showing the decoded word on the screen either in green if correctly classified, or in red if wrongly classified. See supplementary video 1 for an example of the participant performing the online task.

### Error trials

Trials were the participants accidentally spoke during the internal speech part (3) or said the wrong word during the vocalized speech part (20) were removed from all analysis.

#### Neural Firing Rates

Firing rates of sorted units were computed as the number of spikes occurring in 50ms bins, divided by the bin width, and smoothed using a Gaussian filter with kernel width of 50ms to form an estimate of the instantaneous firing rates (spikes/sec).

### Quantification and Statistical Analysis

Analyses were performed using MATLAB R2020b and Python, version 3.8.11.

#### Linear regression analysis

To identify units exhibiting selective firing rate patterns (or tuning) for each of the eight words, linear regression analysis was performed in two different ways: 1) step by step in 50ms time bins to allow assessing changes in neuronal tuning over the entire trial duration; 2) averaging the firing rate in each task phase to compare tuning between phases. The model returns a fit that estimates the firing rate of a unit based on the following variables:

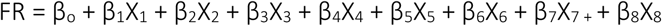

Where FR corresponds to the firing rate of the unit, β_o_ to the offset term which was the average ITI data of the unit, and β corresponds to the estimated regression coefficients (see (Wandelt et al. 2022)).

In this model, β symbolizes the change of firing rate from baseline for each word. A student’s t –test was performed to test the hypothesis of β = 0. A follow up analysis was performed to adjust for false discovery rate between the p-values (Benjamini and Hochberg 1995) (Benjamini and Yekutieli 2001). A unit was defined as tuned if the hypothesis could be rejected (adjusted p value < 0.05, t-statistic) for at least one word. This definition allowed for tuning of a unit to zero, one, or multiple words during different time points of the trial. Linear regression was performed for each session day individually. A 95% confidence interval was computed by performing the student’s t-inverse cumulative distribution function over the 10 sessions.

#### Classification

Using the neuronal firing rates recorded during the tasks, a classifier was used to evaluate how well the set of words could be differentiated during each phase. Classifiers were trained using averaged firing rates over each task phase. A model for each phase was build using linear discriminant analysis (LDA), assuming an identical covariance matrix for each word, which resulted in best classification accuracies. Principal component analysis (PCA) was applied on the training data and PCs explaining more than 95% of the variance were selected as features, and applied to the testing set. Leave-one-out cross-validation was performed to estimate decoding performance. A 95% confidence interval was computed as described above.

#### Cross-phase classification

To estimate shared neural representations between different task phases, we performed cross-phase classification. The process consisted in training a classification model (as described above) on one of the task phases (e.g. ITI) and to test it on the ITI, cue, imagined speech, and vocalized speech phases. The method was repeated for each of the 10 sessions individually, and a 95% confidence interval of the mean was computed. Significant differences in classification accuracies between phases decoded with the same model were evaluated using a paired t-test, with alpha = 0.001.

#### English/Spanish task classification

To evaluate if semantic or phonetic processes are encoded within SMG, data from an English and a Spanish version of the task were merged. Multiunit activity from 96 channels was combined over three session days, resulting in 24 trials per condition. A multiclass support vector machine (SVM) algorithm with radial basis function kernel was used to train a model on one task phase (e.g. ITI), and to test it on the ITI, cue, imagined speech, and speech phases. 10-fold cross validation was performed for model evaluation. A confusion matrix was computed showing classification accuracies per word.

#### Classification performance significance testing

To assess the significance of classification performance, a null dataset was created by repeating classification 100 times with shuffled labels. Then, different percentile levels of this null distribution were computed and compared to the mean of the actual data. Mean classification performances higher than the 95^th^ percentile were denoted with a * symbol and higher than 99^th^ percentile were denoted with **.

#### Demixed principal component (dPCA) analysis

dPCA analysis was performed to break down the activity of the neuronal population into individual components, also called marginalizations, and to observe the explained variance contained in the data for each marginalization. This analysis showed the contribution of each signal variable in the observed data. We followed the method and code (https://github.com/machenslab/dPCA) described in (Kobak et al. 2016), and adapted it to our dataset.

Our dataset had three parameters: timing, cue modality, (e.g. auditory or visual), and word (8 different words). As in the original manuscript, data were decomposed into five parts: condition-independent, cue modality-dependent, word-dependent, dependent on the cue modality-word interaction, and noise. Similar to the covariance decomposition done in ANOVA, individual terms were given by a series of averages. Some of the terms were grouped together as they were individually of lesser interest for this analysis. The “Timing” marginalization combined the time, and cue modality – word – time interaction. The “Cue Modality” marginalization combined the cue modality, cue modality-time interaction, and cue modality-word interaction. The “Word” marginalization combined the word and word-time interaction. To avoid overfitting, a regularization term lambda was used. The analysis was performed as described in (Kobak et al. 2016).

## Supplementary Figures

**Figure S1.**
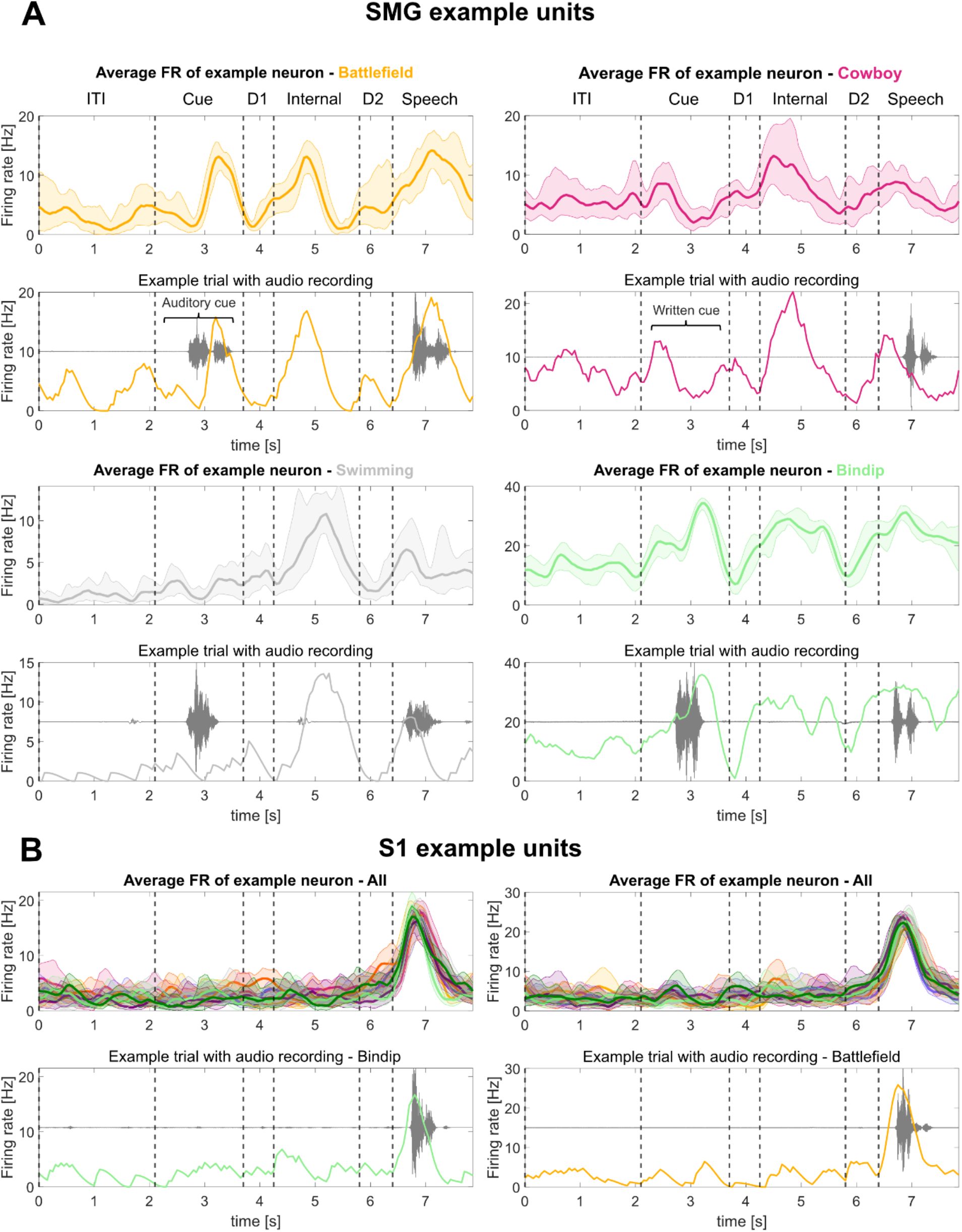
SMG shows firing rate modulation during cue, internal speech, and vocalized speech, S1 shows firing rate modulation during vocalized speech. **A)** Additional example smoothed firing rates of neurons tuned to four words in SMG during the “Auditory cue” and the “Written cue” task. The top part of each word figure shows the average firing rate over eight trials (solid line: mean, shaded area: 95% bootstrapped confidence interval). The bottom part of each figure shows an example trial with associated audio amplitude (gray). Vertically dashed lines indicate the beginning of each phase. **B)** Example smoothed firing rates for S1 over task duration. Tuning of a neuron to all words simultaneously was shown to emphasize generalized speech activity to vocalized words.

**Figure S2.**
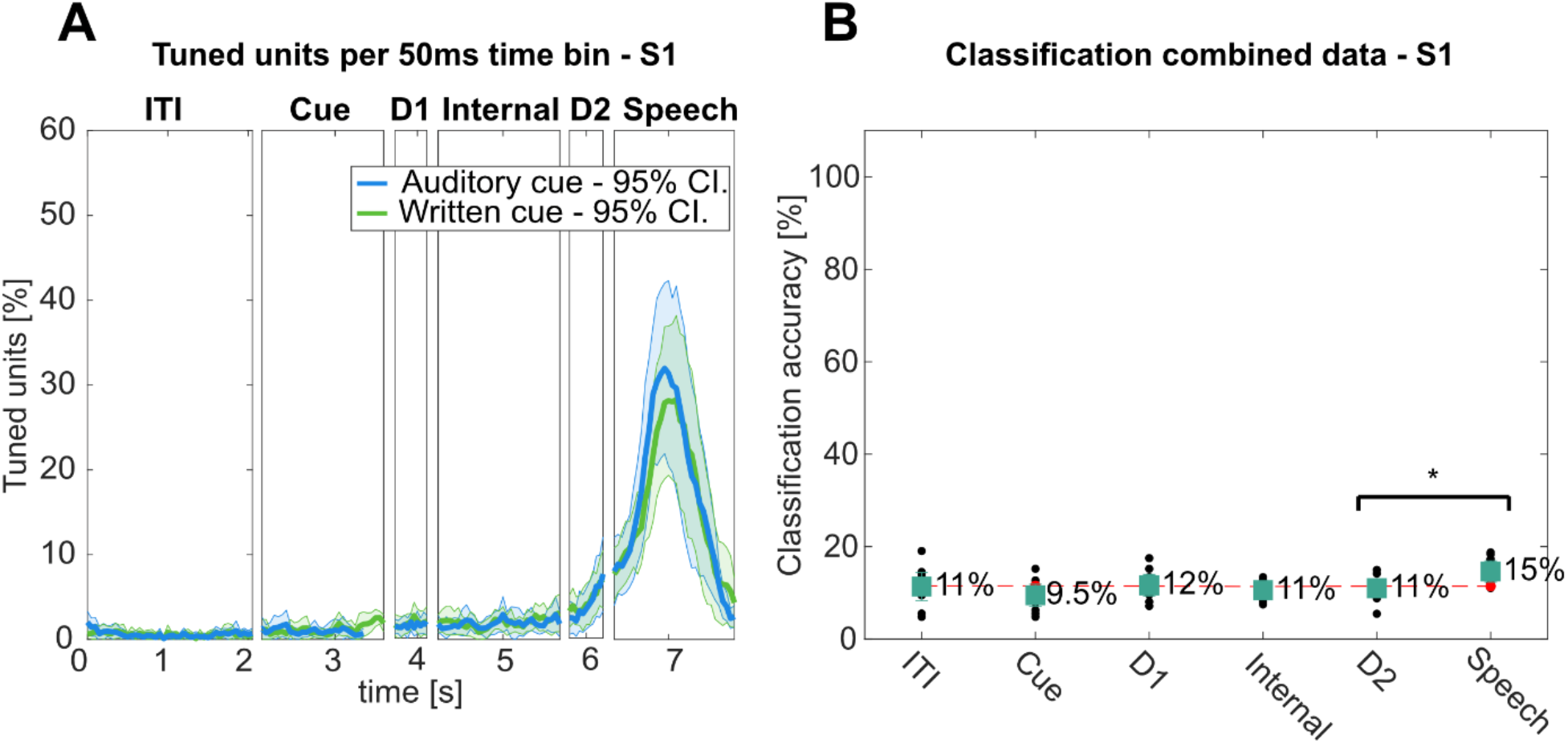
S1 shows generalized word activity during vocalized speech. **A)** Average percentage of tuned neurons to words in 50ms time bins in S1 over the trial duration for “Auditory cue” (blue) and “Written cue” (green) tasks (solid line: mean over 10 sessions, shaded area: 95% confidence interval). **B)** “Auditory cue” and “Written cue” tasks data were combined for each individual session day, and leave one out cross-validation was performed (black dots). PCA was performed on the training data, a LDA model was constructed, and results were plotted with 95% c.i, of the session means. Significance of classification accuracies was evaluated by comparing results to a shuffled distribution (averaged shuffle results = red dots). No classification accuracy was significant. However, classification accuracy during vocalized speech was significantly higher than during the previous delay period (t-test: *p < 0.05). These results show while lip and face activity are represented in putative arm area in S1, it is not significantly decodable.

**Figure S3.**
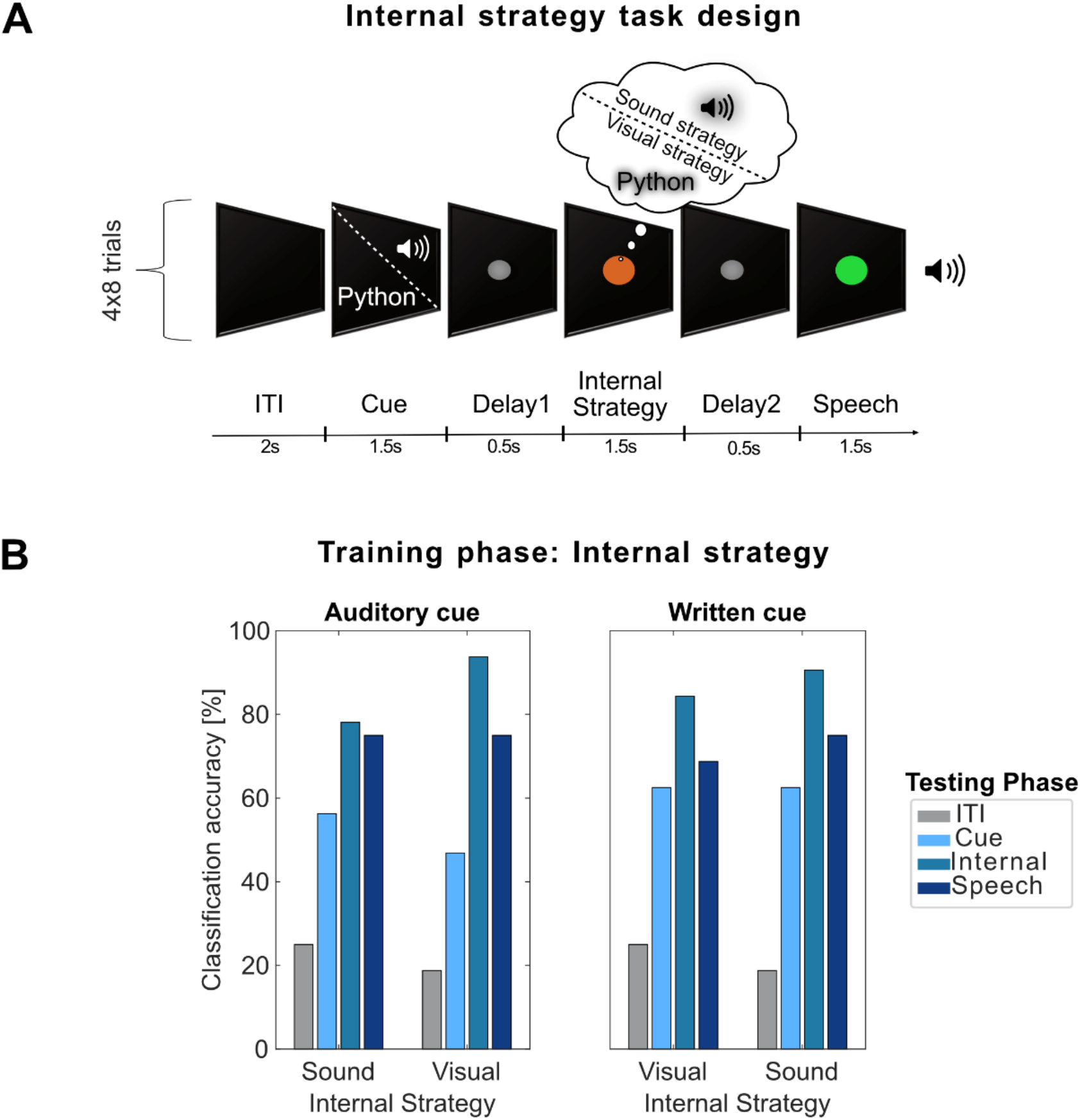
Different internal speech strategies are represented in SMG. **A)** The task was designed to vary the internal strategy the participant was performing during the internal speech phase. Two internal speech strategies were tested: a sound imagination and a visual imagination strategy. For the “sound imagination” strategy, the participant was instructed to imagine the sound of the word. For the “visual imagination” strategy, the participant was instructed to perform mental visualization of the written word. To test if the cue modality (auditory or written) could influence the internal strategy, each internal strategy was run once with an auditory cue, and once with a written cue, resulting in four different task versions (Auditory/Sound, Auditory/Visual, Written/Sound, Written/Visual – see methods). A subset of four words was used for this experiment. **B)** Cross-phase classification was performed by training the model on a subset of data from one phase (e.g. Cue) and applying it on a subset of data from each phase. This analysis was performed separately for each phase, and for each of the four task versions. Plotted here are the results when training on the internal speech phase, and evaluating it on ITI, Cue, Internal, and Speech phases. High classification accuracies (up to 94%) while performing the internal strategy were achieved using both visual and sound imagination strategy.

**Figure S4.**
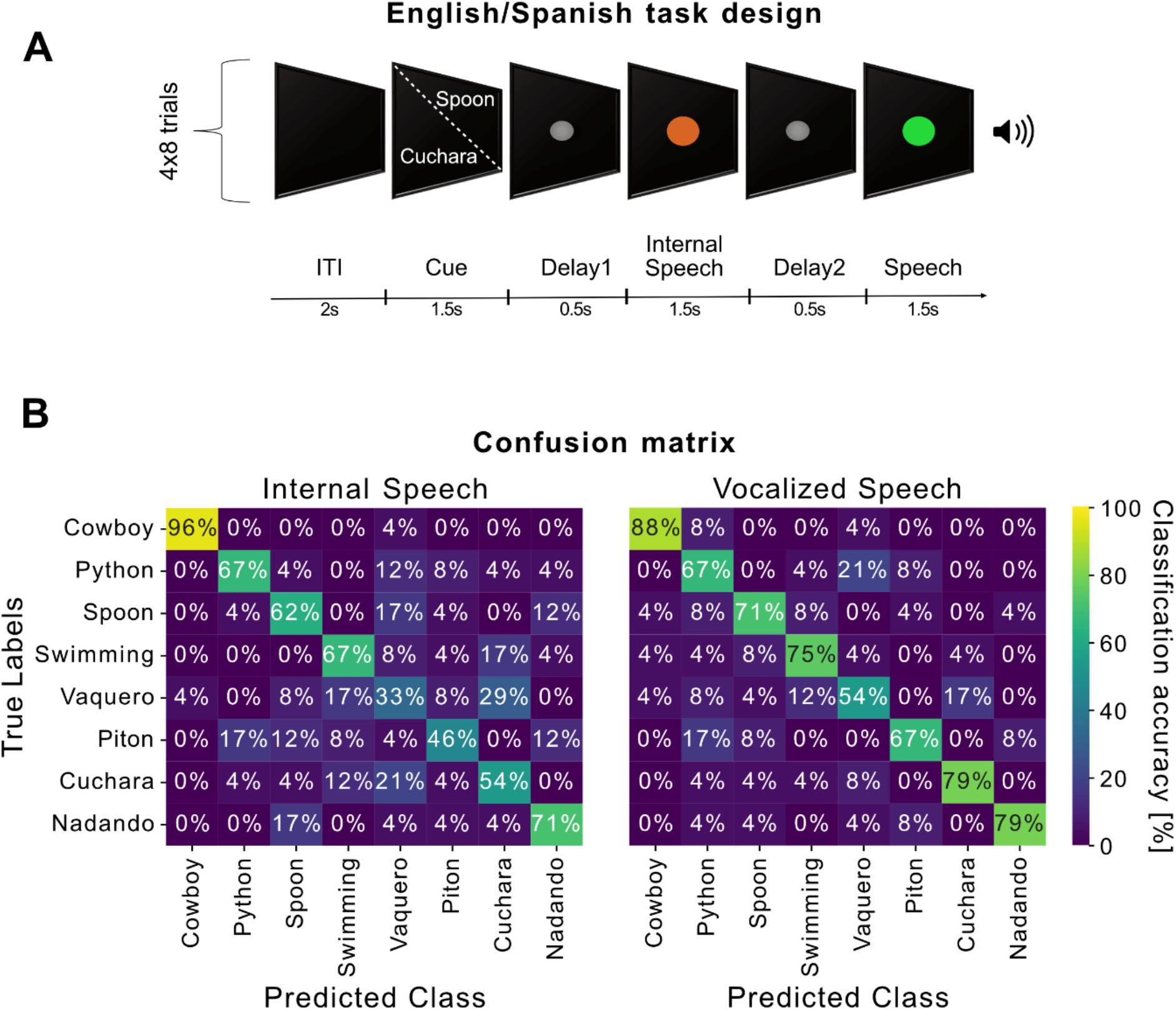
SMG encodes sound phonetics. **A)** The task was designed to probe if SMG encodes stronger the semantics (e.g. the concept of a word) or the phonetics of a word. The bilingual Spanish participant translated four English words into their Spanish counterparts, to ensure the semantic meaning of the word remained the same. The Spanish version of the task was composed of the translated words. On three session days, an English version and a Spanish version of the task were run, resulting in 24 trials per word. The task design remained identical to task 1. **B)** Data was concatenated over session days and languages. A support-vector class model was trained and evaluated with 8-fold cross validation on multiunit channel activity. The confusion matrix during the internal speech phase (left) and vocalized speech phase (right) was calculated. Results indicate the classifier rarely got confused between words with the same meaning, suggesting SMG processes phonetics stronger than semantics both during internal and vocalized speech.

## Notes

### Competing Interest Statement

The authors have declared no competing interest.

### Clinical Trial

NCT01964261

### Author Declarations

Internal Review Board of California Institute of Technology gave ethical approval for this work on 7/25/2022.

